# Immunity to COVID-19 in India through vaccination and natural infection

**DOI:** 10.1101/2021.11.08.21266055

**Authors:** Tresa Rani Sarraf, Shreyasi Maity, Arjun Ghosh, Suchandan Bhattacharjee, Arijit Pani, Kaushik Saha, Dhrubajoyti Chattopadhyay, Gourisankar Ghosh, Malini Sen

**Affiliations:** Indian Institute of Chemical Biology, 4 Subodh Chandra Mullick Road, Kolkata 700064; Biobharati Life Science, EN-35, Sector V, Kolkata 700091; University of California, San Diego, 9500 Gilman Drive, La Jolla, CA 92093; Sister Nivedita University, DG 1/2 New Town, Kolkata 700156

**Author notes:** These authors contributed equally (Tresa Rani Sarraf, Shreyasi Maity, Arjun Ghosh).

## Abstract

In India, <u>Co</u>rona <u>Vi</u>rus-2 <u>D</u>isease-2019 (COVID-19) continues to this day, although with subdued intensity, following two major waves of viral infection. Despite ongoing vaccination drives to curb the spread of COVID-19, the potential of the administered vaccines to render immune protection to the general population, and how this compares with the immune potential of natural infection remain unclear. In this study we examined correlates of immune protection (humoral and cell mediated) induced by the two vaccines Covishield and Covaxin, in individuals living in and around Kolkata, India. Additionally, we compared the vaccination induced immune response profile with that of natural infection, evaluating thereby if individuals infected during the first wave retained virus specific immunity. Our results indicate that while Covaxin generates better cell-mediated immunity toward the Delta variant of SARS-CoV-2 than Covishield, Covishield is more effective than Covaxin in inducing humoral immunity. Both Covishield and Covaxin, however, are more effective toward the wild type virus than the Delta variant. Moreover, the overall immune response resulting from natural infection in and around Kolkata is not only to a certain degree better than that generated by vaccination, especially in the case of the Delta variant, but cell mediated immunity to SARS-CoV-2 also lasts for at least ten months after the viral infection.

## Introduction

COVID-19 (<u>Co</u>rona <u>Vi</u>rus <u>D</u>isease-2019) is caused by infection with the <u>S</u>evere <u>A</u>cquired <u>R</u>espiratory <u>S</u>yndrome causing <u>Co</u>rona <u>V</u>irus – 2 (SARS-CoV-2), a single strand RNA virus [1; 2]. The disease is believed to have originated in China in December 2019 [3; 4]. Since that time, COVID-19 has ravaged across several countries causing many fatalities. The World Health Organization (WHO) designated COVID-19 as a pandemic in March 2020 [1]. So far, there have been multiple major SARS-CoV-2 associated infection waves worldwide. India suffered from two such waves the first one spanning July-November, 2020, and the second one spanning April – June, 2021.

SARS-CoV-2 enters into the human host usually through the upper respiratory tract and binds to the ACE2 receptor expressed on host epithelial cells through its <u>S</u>pike protein - <u>R</u>eceptor <u>B</u>inding <u>D</u>omain, the S-RBD [2; 5]. The S-RBD – ACE2 interaction disrupts the normal function of ACE2, which is essential for the maintenance of proper human physiology [6]. The ensuing reactions to the establishment of the viral infection in the human host range from standard inflammation causing mild disease to uncontrolled cytokine storm, which may even lead to death [7; 8]. Several studies have indicated that blockade of the activity of the S-RBD domain of the virus would be effective in restricting disease progress. These studies have paved the way toward the development of several vaccines against the Spike protein of the virus, an example being the Oxford– AstraZeneca chimpanzee adenovirus vectored vaccine ChAdOx1 nCoV-19 (AZD1222) [9]. The same vaccine is known as Covishield in India. Conventional vaccines against the whole virus have also been developed to fight off infection, Covaxin (product of Bharat-Biotech, India) being an example [10].

SARS-CoV-2 infection has resulted in many hospitalizations and deaths in India. With the dipping of the first COVID wave, a second wave, which emerged with few mutant forms of the SARS-CoV-2 virus, has led to even more damage than the first. A major mutant (DELTA variant) with L452R and T478K double mutations in the S-RBD domain has dominated the second infection wave [11; 12]. With the fag end of this wave still continuing, danger of a third wave looms. Although a major vaccination drive has begun, only about 30% of the massive Indian population has so far been fully vaccinated (two vaccine doses). Moreover, even among the sparsely vaccinated fraction, there have been few reports of COVID-19 related deaths. While several studies targeted numerous vaccines to determine their efficacy against several SARS-CoV-2 strains [13], very few comparative studies have been done on the two vaccines most used in India, especially with respect to natural infections mediated by wild type virus (Wuhan-HU-1 isolate) and the DELTA variant. In this scenario, it is very important to evaluate and compare the efficacy of the currently administered vaccines, Covishield and Covaxin, particularly with reference to the immune protection generated by natural infections. To this end, we undertook a comprehensive study aimed at assessing the immune response profiles prevalent in the general Indian population of West Bengal, using blood samples from different cohorts: (i) the vaccinated with Covishield or Covaxin, (ii) the naturally infected during the first and second waves of infection and (iii) unvaccinated apparently healthy controls (reference group). Our goal was to evaluate the levels of immune response in the different study groups, and thereby analyze the immune protection generated through vaccination and natural infection.

In a prior population based epidemiological study centered on West Bengal we had demonstrated about 90% effectiveness of the Covishield vaccine in generating antibody response (humoral immunity) against wild type SARS-CoV-2 [14]. Here, we extended our study to compare the effectiveness of the Covishield and Covaxin vaccines in terms of both humoral (with virus neutralization potential) and cell mediated immunity to the wild type and mutant (DELTA) variant(s) of SARS-CoV-2. Moreover, we evaluated the efficacy of both the vaccines in relation to the potential immune protection rendered by natural infection.

## Methods

### Human Subjects & Ethical Declaration

An Institutional Review Board (IRB) on Human Subjects was constituted following the guidelines of the Indian Council for Medical Research (ICMR). The IRB read and discussed the research proposal, and provided a certificate of approval dated December 20 2020, for the research conducted in this study. Samples were collected from donors following the protocol approved by the IRB after receiving their signed consent. Gram Panchayat (Village Council) leaders along with the local non-government organizations helped assemble donors in the rural places and were present during sample collection. In Kolkata, donor groups were assembled by various non-government organizations.

### Isolation of blood plasma and PBMC

Plasma and PBMC isolation was performed following published protocols, with minor modifications [15; 16]. Approximately 10ml blood collected in a vial (BD Vacutainer Cat. No. 368856) from each donor was centrifuged at 200g to separate the plasma. The plasma, collected very carefully in a separate centrifuge tube was again centrifuged at 1000g to separate platelets. After removing platelets, plasma was stored in −80°C until used for antibody measurement. Following plasma separation blood was diluted in 1:1 ratio with 1XPBS for Histopaque density gradient centrifugation at 350g without brakes for 20min. Subsequently, the buffy coat (comprising PBMC), which was collected in a separate tube was washed 2 times with 1X PBS (centrifuged at 350g for 8 min each time after suspension) to remove residual histopaque. To remove platelets a last wash was done at 200xg for 10 min. PBMC pellet was resuspended in 1ml RPMI for haemocytometer counting. Following centrifugation again at 350g for 8 min, isolated PBMC were frozen by resuspension in cold FBS with 10% DMSO. 6x 10^6^ cells/ml were aliquoted in each cryovial and immediately stored in Mr. Frosty freezing container in −80°C. After 24hr cryovials were transferred from −80°C to liquid nitrogen until used for cell mediated immune response experiments.

In some cases, blood sera were used instead of plasma. 1 mL of blood was collected from each subject, followed by an overnight incubation at 4 °C and centrifugation for 10 min at 1000Xg. Sera were collected in fresh tubes and saved at −80 °C until further use.

### Measurement of total Spike protein specific antibodies (IgG/IgM/IgA)

ELISA was performed following the method published by Stadlbauer D et.al [17] with some modifications using a test kit developed in-house. Briefly, 100 ng of antigen (same for S-RBD and Delta variant) was coated in each well of 96 well high binding ELISA plates (Corning, USA). The antigen was incubated for 16 h at 4 °C, followed by washing with 1X TBST. The antigen coated plates were then blocked with 5% non-fat dry milk (Himedia, India) dissolved in TBST and kept for 1h at room temperature. Then the plates were washed with 1X TBST before addition of sera (plasma) at a 1:100 dilution. Incubation with plasma was done for 90 minutes at room temperature before washing with 1X TBST. The secondary antibody (anti mouse IgG/IgM/IgA-HRP conjugated, Sigma, USA) in a 1:5000 dilution was then added to each well and incubation was done at room temperature for 45 minutes. After washing with 1X TBST, the plates were developed according to the standard TMB method and OD was measured at 450 nm on a BioRad Mark microplate reader.

### Estimation of total (IgG/IgA) neutralizing antibody

Briefly, 100 ng of antigen (S-RBD; same for WT and DELTA variant) was coated on each of 96 wells of high binding ELISA plates (Corning, USA). The plates were incubated for 16 h at 4 °C followed by washing with 1X TBST. The antigen coated plates were then blocked with 5% non-fat dry milk (Himedia, India) dissolved in TBST and kept for 1h at room temperature. Following washing with 1X TBST, a 100 uL mix of 1:10 diluted plasma (or sera) with 20 ng of human angiotensin converting enzyme-2 (ACE-2) was added to each well and incubation was continued for 90 minutes at room temperature before washing again 4 times with 1X TBST. Since human ACE-2 is tagged with mouse Fc for its binding to anti-mouse secondary antibody, the secondary antibody (anti mouse IgG-HRP conjugated, Sigma USA) was added in 1:5000 dilution to the wells. Following 45 min incubation at room temperature and subsequent washing with 1X TBST, the plates were developed according to standard TMB method and OD was measured at 450 nm in BioRad Mark microplate reader five mins after the reaction was stopped (reading A). Since mouse secondary antibody also cross reacts with human antibody to a certain extent, a control experiment was needed for background correction. Thus a parallel ELISA was carried out identically except that ACE2 was omitted (reading B). In another reaction, 20 ng ACE2 in 100 uL 1XTBST was incubated with each RBD coated well followed by washing, secondary antibody incubation and development (reading C). OD_450_ in C represents the maximum binding of 20 ng ACE2 to coated RBD. OD_450_ in A represents ACE2 remaining after plasma/sera competition. OD_450_ in B represents cross-reactivity of human antibody to mouse secondary antibody. Percent neutralization was calculated using the formula [100-(A-B)/C X 100]. All reactions were done in duplicates, and some were triplicates.

### Ni-NTA affinity pulldown assay to test binding and neutralization

To visualize specific binding competition, we used the Ni2+ pull down assay. In this case, 400 ng poly-histidine fused RBD-WT or RBD-DELTA was bound to N2+ NTA beads (10 uL) and incubated with positive or negative sera (500 uL 1:10 dilution) as determined by ELISA or ACE-2-Fc (400 ng) in a buffer containing 150 mM NaCl, 1% Tween20, 25 mM Tris-HCl 7.5. Binding was allowed to occur for 1 hour, followed by extensive washing with the same buffer (500 uL) 4 times. The bead after washing was treated with 20 uL of 2X SDS loading buffer, followed by separation of the bound protein by SDS-PAGE and staining with Coommassie Blue. The competition binding was done identically except both ACE-2 and plasma were mixed with ACE-2 bound beads. Control reactions were also done where RBD was eliminated in the binding reaction.

### Estimation of cell mediated immunity

Cell mediated immune response was evaluated following published protocols [15; 16; 18]. For all assays cells were thawed in prewarmed water (37°C) and diluted in 10ml prewarmed RPMI media (10% FBS, 1% Penicillin, Streptomycin, 1% l-glutamine). Following brief centrifugation, cells were cultured in 10ml RPMI media for 18hrs under normal tissue culture conditions and subsequently counted using heamocytometer. Subsequently, cells were plated in 24 well plates at 2.4x 10^5^ cells /well and incubated with 2μg/ml of either the wild type or the mutant version (L452R-SRBD) of SARS-Cov-2 S-RBD for 24hr, with Brefeldin added into the culture medium for the last 4hrs. For each protein batch, just the protein diluent (PBS with 50% glycerol) was used as the vehicle control. Protein incubation was followed by one wash with 1XPBS, fixation with 1% paraformaldehyde for 10min at room temperature, 3 washes in 1XPBS, and storage at 4°C overnight. Next day PBMC were permeabilized with 0.1% Tween 20/PBS for 10min at room temperature, washed in permeabilization buffer and stained with Anti human CD4 (PE, Biolegend), Anti human CD8 (APC, Biolegend), Anti human IFN*γ* (PE-CY7, Biolegend) and Anti human CD40L/CD154 (Alexa fluor 700, Biolegend) for 1hr at 4°C. After washing once each with permeabilization buffer and 1XPBS, PBMC were resuspended in 1XPBS for acquisition on BD LSRFortessa Cell Analyzer and analyzed using FCS Express 5 software.

### Data analyses

The cutoff was determined using sera from 25 healthy controls known to be uninfected from our previous study. The mean of the OD values of the negative control replicate plates plus three times the standard deviation of the OD value distribution gave the cutoff. 95% confidence intervals were calculated by using the formula 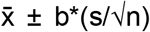, where 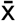 is the sample mean, b is z-score when sample number is greater than 30 or t-score when sample number is less than 30, s is standard deviation, and n is sample number. *P*-values between pre-COVID and post-COVID samples were calculated by one sample one tailed t-test. Briefly, t-statistic was calculated by using the formula 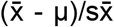, where 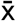 is sample mean, μ is negative control (population) mean, and 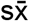 is s/√n (s is sample standard deviation and n is sample number). Then t-table was used to determine the *p*-value range from the t-statistic. iMFI for cell mediated immune response was calculated by multiplying the percent of activated cells with the corresponding geometric mean fluorescence intensity. Samples were analyzed by distribution plot with Graph Pad Prism version 8.3.1 software.

## Results

### Experimental strategy

We strategized an investigative plan for substantiating a prior study focused on SARS-CoV-2 infection rate and vaccination efficacy in West Bengal, India, and extending it further for comparing the extent of immune response to SARS-CoV-2 infection and vaccination. Earlier, we had reported that the infection rate during the first infection wave with the original SARS-CoV-2, which we refer to here as the wild type (WT) virus, in the population in and around Kolkata, India was roughly 40% [14]. Subsequently, the second wave of infection, mediated mostly by the DELTA variant, hit the city’s population starting around the end of March 2021 and lasted till the end of June. By the end of July 2021, a substantial fraction of the city’s population received two doses of vaccines approved in India, - Covishield (same as the Oxford-Astrazeneca) and Covaxin (originated in India). In this study we focused on measuring the neutralizing antibody (humoral immume response) and T cell mediated immune response of the naturally infected (both first wave and second wave) and vaccinated population. To this end, we organized five experimental groups (Table 1). The first two groups comprised the population vaccinated with either Covishield (1), or Covaxin (2). These two groups had no known history of infection, but some of them could have been naturally infected without any symptoms. Groups 3 and 4 were naturally infected either during the first wave (3) or during the second wave (4). Group3 had developed antibodies against S-RBD (WT) by the end of December 2020 but were mostly asymptomatic [14]. Their blood samples were collected again in July 2021 for the current study. Individuals in Group 4 were infected during May-June 2021. All Group 4 individuals were symptomatic, and most were hospitalized. Blood samples were collected from them post recovery. The last group (Group 5) served as a reference or control. The subjects in Group 5 had no known history of infection and were not vaccinated. However, some of them could have been infected asymptomatically.

**Table 1:** Summary of test subjects for five different groups and their humoral immunity

| Group | Total | Male | Female | Days After<br>Vaccination/<br>or infection<br>(mean) | Seropositive<br>WT/DELTA (%) | Neutralization<br>positive<br>WT/DELTA<br>(%) |
| --- | --- | --- | --- | --- | --- | --- |
| Covishield | 149 | 96 | 53 | 88 | 87/86 | 94/86 |
| Covaxin | 42 | 17 | 25 | 77 | 81/79 | 86/74 |
| 1<sup>nd</sup> Wave | 16 | 4 | 12 | 275 | 82/60 | 75/47 |
| 2<sup>st</sup> Wave | 14 | 7 | 7 | 71 | 86/86 | 100/93 |
| Healthy<br>Control | 19 | 13 | 6 |  | 47/40 | 23/7 |

In order to estimate humoral and cell mediated immunity to SARS-CoV-2 generated through vaccination and natural infection, blood samples were collected from all designated groups as explained in Table 1. In all the samples, the titer of anti-S-RBD antibody against both the WT and DELTA variant specific S-RBD proteins and its potential to neutralize ACE2-S-RBD interaction were evaluated. In essence, we determined the total antibody (IgG/IgM/IgA) titer against the antigens S-RBD WT and S-RBD DELTA (hereafter referred to as RBD-WT and RBD-DELTA, respectively) and the ability of these S-specific antibodies to replace the host receptor protein, ACE2, from RBD-WT or RBD-DELTA by virtue of antigen-antibody interaction. Furthermore, we analyzed the T cell response toward RBD-WT and RBD-DELTA within a subgroup (Supplementary Table 1) of each group of samples by quantifying the IFN*γ* and CD40L levels of CD4 and CD8 T cells as a measure of cell mediated immunity.

### Estimation of humoral immunity to SARS-CoV-2 elicited through vaccination and natural infection

#### Levels of antibody detecting RBD-WT and RBD-DELTA among the test groups

We determined the antibody titer in all the plasma samples under study by ELISA. First, we determined the cut off OD values for both RBD-WT and RBD-DELTA using sera that were previously shown to be seronegative [14]. The cut off was found to be 0.25 for both WT and DELTA (Supplementary Figure 1A-C). Figure 1A and 1B show the sero-sensitivity for all samples tested. As described in Figure 1A, corroborating our previous finding [14], Covishield showed about 90% efficacy in generating anti-RBD-WT antibodies, i.e. about 90% of the volunteers tested turned out seropositive after vaccination. Using similar assay conditions, Covaxin, however, showed about 6% less efficacy than Covishield. Both the first and second infection waves were almost as effective as the two vaccines in producing anti-wild type S-RBD antibody in the infected population, with about 82% (first wave) and about 86% (second wave) of the volunteers showing seropositivity. The antibody profile of the same cohorts against the RBD-DELTA protein was quite similar (Figure 1B). It should be noted that all first wave samples were seropositive against RBD-WT in January 2021. As shown in Supplementary Figure 1D, nearly all samples showed waning S-specific antibody with a couple even below the cut off. This is not surprising since these subjects were infected anytime between July and October of 2020, about 8-11 months before the time of the second collection.

**Figure 1:**
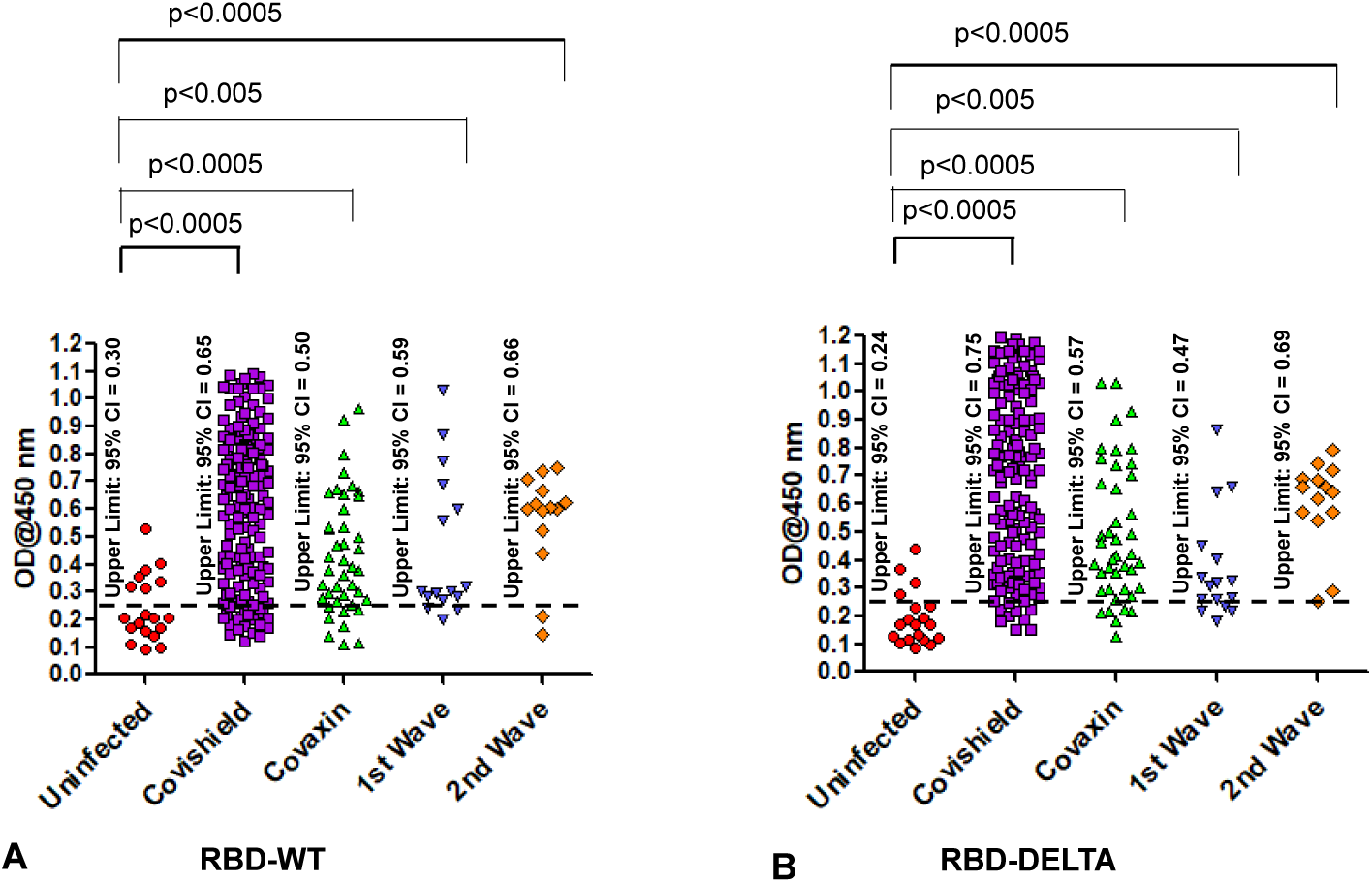
Seroreactivity of infected and vaccinated population against Spike (S)-RBD. A. Seroreactivity against RBD-WT of all samples in each of the five groups. 95% confidence intervals and p values for comparing individual distributions to that healthy control are shown. B. Seroreactivity against RBD-DELTA of all samples in each of the five groups. 95% confidence intervals and p values for comparing individual distributions to that healthy control are shown.

#### Levels of antibody neutralizing RBD-WT and RBD-DELTA binding with ACE2 among the test groups

In order to test the ability of anti-RBD IgG to neutralize ACE2-RBD interaction, we first quantified the efficiency of RBD-WT/RBD-DELTA-ACE2 interaction, by ELISA. Indeed, ACE2 bound to both RBDs with nearly equal efficiencies, with 100% binding saturation at ∼50 ng ACE2 (Supplementary Figure 2A). ACE2-RBD neutralizing potential of a plasma sample was evaluated by adding a mixture of ACE2 (25 ng) and test plasma to either WT- or DELTA-RBD coated wells (explained in the method section). Upon examining the neutralizing potential of the plasma samples by ELISA, we found that not all were 100% effective in blocking the ACE2:RBD interaction *in vitro*. We observed high neutralization efficiency (95% confidence interval: 51.7-63.4) for Covishield and second wave samples (95% CI: 33.6-67.6) and low for Covaxin (95% CI: 27.5-45.5) and the first wave samples (95% CI: 2.9-33.5) against the WT RBD (Figure 2A). For the DELTA variant, all the tested cohorts excepting the 2^nd^ wave samples produced slightly lower detectable levels of neutralizing antibody but maintained the same ranks in the neutralization efficiency (Figure 2B). Quite interestingly, few of the samples collected from the uninfected and unvaccinated donors also showed some degree of seropositivity and neutralization potential, although at a much lower level than the rest of the cohorts, implying the presence of cross-reactive antibodies against S-RBD overlapping antigenic epitopes [19; 20].

**Figure 2:**
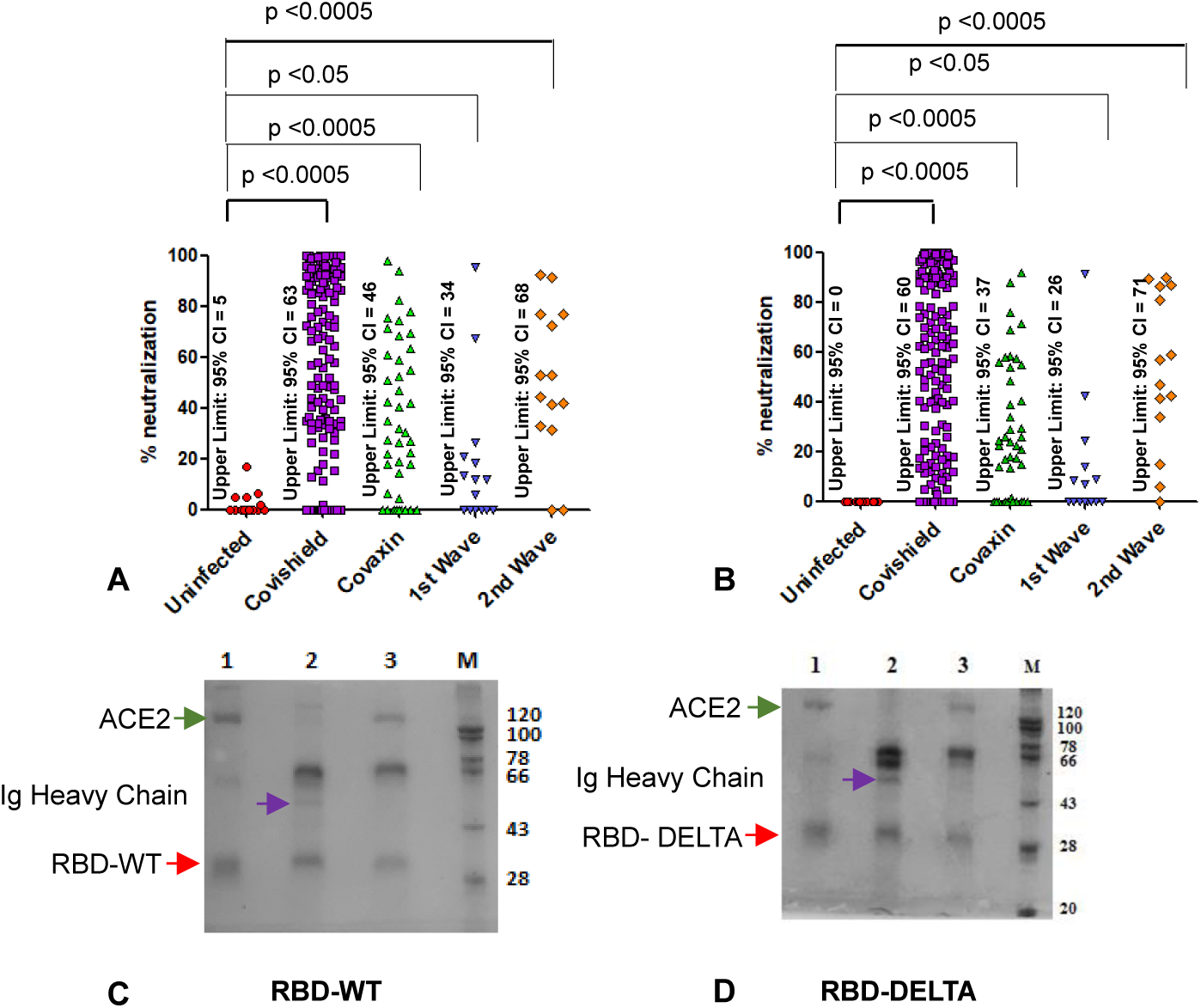
Competition between plasma/sera from infected and vaccinated people and ACE2 for RBD binding as surrogate neutralization activity. A: Neutralization as percent signal inhibition by sera/plasma samples (1:10 dilution) from five different groups with 100 ng His-RBD-WT as the capture antigen in competition with 20 ng ACE2 by ELISA. Each sample was run in duplicates and each dot represents a mean of two readings. B: Neutralization as percent signal inhibition by the same sera/plasma samples as in A with 100 ng His-RBD-DELTA variant as the capture antigen in competition with 20 ng ACE2. Each sample was run in duplicates and each dot represents a mean of two readings. Competition assays of each sample for both RBD-WT and RBD-DELTA were done in parallel in same plate for better reproducibility. C. Competition was tested using the His-Ni2+ affinity pulldown assay. His-RBD-WT (400 ng) captured by Ni2+ affinity beads (red arrow) bound pure ACE2 (green arrow, 400 ng) (lane 1). A sample with RBD-specific antibody efficiently competed ACE2 binding (lane2) but not a sample negative for RBD-specific antibody (lane 3). M denotes MW standards. ‘violet arrow’ denotes Ig-heavy chain that specifically bound to RBD-WT. D. Same as C except RBD-DELTA was used as captured antigen. Positive and negative plasma used were same as in C.

To further corroborate the presence of the RBD-WT and RBD-DELTA specific neutralizing antibodies, we demonstrated both interaction between RBD and ACE2, and competition between ACE2 and specific antibodies for RBD binding. Herein, we performed the affinity pull-down assay followed by SDS-PAGE analysis. Since both RBD-WT and RBD-DELTA used for this assay are poly-histidine tagged, the proteins were initially bound to Ni-NTA beads, which were then subjected to treatment with either anti-S-RBD positive or negative plasma or pure ACE2. As shown in Supplementary Figure 2B, RBD-WT-bound Ni-NTA beads but not unbound beads pulled down ACE2. RBD-WT-bound Ni-NTA beads also pulled down Ig from the seropositive sample but not from the seronegative (Supplementary Figure 2C). Accordingly, in the competition assay, when ACE2 was added to the Ni-NTA beads bound to RBD-WT or RBD-DELTA in the presence of plasma containing S protein-specific antibodies we found a drastic reduction in ACE2 binding, with concomitant binding of Ig heavy chain to the beads. As expected, plasma with no S protein-specific antibody failed to compete for ACE2 binding (Figure 2C & D).

Overall, these results suggest that while natural infection with SARS-CoV-2 is as effective as vaccination in generating an antibody response, this humoral immunity may not last long enough to fight off future infections. Whether the currently administered vaccinations will provide a long-term humoral immunity against the virus is a matter that only time will confirm.

### Evaluation of cell mediated immunity to SARS-CoV-2 generated through vaccination and natural infection

T cell activation and cytokine expression being an important facet of immunity [16; 18; 21; 22; 23], we also focused on the level of T cell response to vaccination and natural infection, in the general population of West Bengal. To this end, blood samples collected from the same groups of volunteers in the specified time frame as explained in the previous section were tested for CD8+ and CD4+ T cell immunity against both the WT-RBD and DELTA-RBD proteins. Since IFN*γ* is an established cytokine for estimating both CD8+ and CD4+ T cell activation in response to viral antigens [18; 21], we evaluated the levels of intracellular IFN*γ* in the CD8+ and CD4+ T cells of a subgroup of individuals in each of the 5 groups summarized earlier upon stimulation with either the WT-RBD or the DELTA-RBD protein (Table 1 and Supplementary Table1). As an additional requisite of T cell guided immunity, we also assessed the level of expression of CD40 ligand (CD40L/CD154) on account of its documented role in the clonal expansion of antigen specific CD8+ and CD4+ T cells through interactions with the CD40 receptor expressed on antigen presenting cells [18; 24; 25]. The idea was to estimate recall immune responses generated in cells expected to be primed through vaccination or natural viral infection, after stimulation with the appropriate viral antigen *ex vivo* [16; 26; 27; 28; 29; 30].

Increase in CD8 and CD4 T cell specific IFN*γ* and CD40L levels in response to the S-RBD protein were analyzed by fluorescence activated cell sorting (FACS) of the peripheral blood mononuclear cells (PBMC) harvested from the blood samples with the use of appropriate fluorophore tagged antibodies, as explained in Materials and Methods. The gating strategy for this analysis is illustrated in Figure 3A. In this study, as summarized in Supplementary Table 1, we included 16 individuals each from the two vaccinated groups (Groups 1 & 2), 12 and 9 individuals respectively, from the naturally infected groups (Groups 3 & 4), and 5 individuals, who had undetectable levels of anti-S-RBD neutralizing antibody from the control group (Group 5). Sero-sensitivity and neutralization efficacy of these subjects are shown in Supplementary Figure 3A-D. In the FACS analysis of each PBMC sample of each group, the change in the level of the T cell activation marker (IFN*γ* or CD40L) after stimulation with the S-RBD protein was assessed with respect to the corresponding ‘no protein’ control i.e. vehicle control (vc), as depicted in Figure 3B by specific examples from both the vaccinated and natural infection groups. Since both the percent of activation marker positive cells and the corresponding geometric mean fluorescence intensity are important contributors of immune activation, both values were integrated into an integrated <u>M</u>ean <u>F</u>luorescence Intensity (iMFI: percent positive cells X geometric mean of fluorescence intensity) value as a measure of activation [15; 29]. Percent increase in T cell activation (iMFI) for each marker after protein stimulation was calculated by multiplying [iMFI (S-RBD) – iMFI (vehicle control)] / iMFI (vehicle control) with 100.

**Figure 3:**
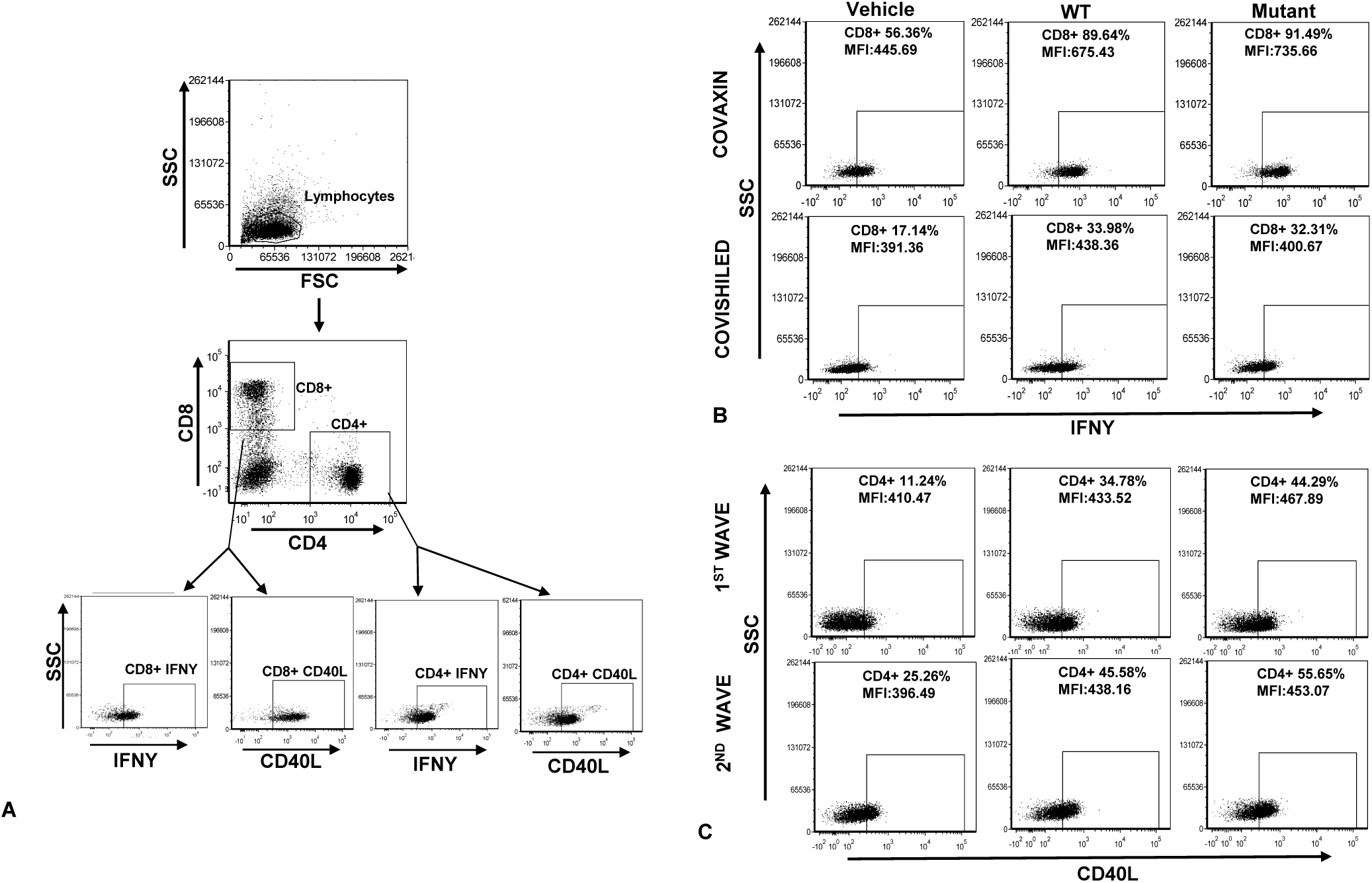
Depiction of FACS on PBMC collected from human blood samples for analysis of cell mediated immunity. **A**: Representation of initial gating on PBMC for CD4+ and CD8+ lymphocytes followed by demarcation of IFN*γ* and CD40L positive populations, based on evaluation of geometric mean fluorescence intensity generated by antibody staining. **B:** Representation of change in percent and geometric mean fluorescence intensity (noted in parenthesis) of IFN*γ* positive CD8T lymphocytes in response to stimulation by SARS-Cov-2 specific S-RBD protein. **C**: Similar representation with respect to CD40L positive T lymphocytes.

#### T cell IFNγ and CD40L specific recall immune response

The values of the T cell IFN*γ* and CD40L specific recall immune response in each of the 5 groups, expressed as mean of the percent increase in iMFI (S-RBD protein vs. vehicle control) in all samples belonging to each group, are summarized in Table 2. We noted that while there was no mean percent increase in iMFI in the control group (Group 5) in response to protein, in the other groups (Groups 1-4), the mean percent increase was distributed through a wide range. Overall, in terms of mean CD8 and CD4 T cell IFN*γ* response to RBD-WT, both the vaccinated groups were comparable to the natural infection groups and to each other. The CD40L specific mean response by and large was lower compared to the IFN*γ* response in all the experimental groups; however, the same trend as that observed for IFN*γ* was also present here. In the case of T cell response to RBD-DELTA, the scenario was distinctly different. Barring the CD8-CD40L response, all other measured T cell responses were several folds lower in the vaccinated groups as compared to the natural infection groups, where the response was even slightly better than that observed in the case of RBD-WT. Nevertheless, although less effective toward the mutant as compared to wild type, Covaxin showed at least 2-3 fold better T cell response toward the mutant than Covishield. Graphical representations of the analyses of T cell IFN*γ* and CD40L response toward RBD-WT and RBD-DELTA in the different groups as iMFI are projected in Supplementary Figures 4 and 5 (Panels A - D).

**Table 2:** Mean of percent increase in iMFI in response to protein (RBD-WT or RBD-DELTA) in test subjects of different groups, 1 through 5.

| Viral Antigen specific immune response |  |  |  |  |  |  |  |  |
| --- | --- | --- | --- | --- | --- | --- | --- | --- |
| Cell Mediated Immunity ( Mean of percent increase in iMFI : Viral protein vs. Vehicle Control) |  |  |  |  |  |  |  |  |
| Groups | CD8-IFNγ (RW) | CD4-IFNγ (RW) | CD8-CD40L (RW) | CD4-CD40L (RW) | CD8-IFNγ (RD) | CD4-IFNγ (RD) | CD8-CD40L (RD) | CD4-CD40L (RD) |
| Covishield | 54.25% | 31.10% | 12.27% | 30.99% | 10.61% | 4.75% | 1.18% | 3.49% |
| Covaxin | 36.38% | 27.20% | 7.18% | 26.68% | 21.11% | 13.57% | 7.02% | 11.98% |
| Infection 1st wave | 41.28% | 23.23% | 10.79% | 44.60% | 50.35% | 25.91% | 6.74% | 54.46% |
| Infection 2nd wave | 53.54% | 41.10% | 2.46% | 27.53% | 81.08% | 40.93% | 0.25% | 33.59% |
| Uninfected | ND | ND | ND | ND | ND | ND | ND | ND |
| RW = RBD-WT |  |  |  |  |  |  |  |  |
| RD = RBD-DELTA |  |  |  |  |  |  |  |  |
| ND = Not Detected |  |  |  |  |  |  |  |  |

Table 3 summarizes the percentage of individuals in each group who displayed a positive T cell IFN*γ* or CD40L recall response to viral antigen. In the vaccinated groups, the percentage of individuals with a positive recall response to RBD-WT was quite similar, with some variation in numbers with respect to IFN*γ* and CD40L. The percentage of T cell responders to RBD-WT in the natural infection groups with respect to IFN*γ* and CD40L was also in the same range. In the case of RBD-DELTA, the overall percentage with a positive T cell recall response was significantly higher in the natural infection groups than in the vaccinated groups. However, as in the case of percent increase in iMFI (Table 2), the percentage of individuals showing T cell response in the Covaxin group was higher than that in the Covishield group. The T cell response seen in 1-2 individuals in the control group could be because of the presence of cross-reactive T cells, which is not an uncommon phenomenon [19].

**Table 3:** Percentage of test subjects positive for cell mediated immunity

| Percentage of individuals with T cell recall response |  |  |  |  |  |  |  |  |
| --- | --- | --- | --- | --- | --- | --- | --- | --- |
| Groups | CD8-IFNγ (RW) | CD4-IFNγ (RW) | CD8-CD40L (RW) | CD4-CD40L (RW) | CD8-IFNγ (RD) | CD4-IFNγ (RD) | CD8-CD40L (RD) | CD4-CD40L (RD) |
| Covishield | 81.25% | 75.00% | 81.25% | 75.00% | 56.25% | 56.25% | 50.00% | 50.00% |
| Covaxin | 93.75% | 93.75% | 75.00% | 62.50% | 68.75% | 62.50% | 62.50% | 50.00% |
| Infection 1st wave | 75.00% | 75.00% | 66.67% | 66.67% | 66.67% | 75.00% | 66.67% | 75.00% |
| Infection 2nd wave | 88.89% | 88.89% | 66.67% | 77.78% | 88.89% | 88.89% | 44.44% | 55.56% |
| Uninfected | 20.00% | 20.00% | 40.00% | 40.00% | 40.00% | 40.00% | 20.00% | 20.00% |
| RW = RBD-WT |  |  |  |  |  |  |  |  |
| RD = RBD-DELTA |  |  |  |  |  |  |  |  |

Taken together, it appears that Covaxin generates better T cell immunity than Covishield against infections with the mutant virus. However, natural infection from SARS-CoV-2 is a better driver of virus specific cell mediated immune response than both vaccines. Additionally, T cell immunity from natural infection seems to last at least for as long as ten months. Whether such immunity can be generated through vaccination remains to be tested.

## Discussion

Escalating fatalities from worldwide dissemination of SARS-CoV-2 infection and designation of COVID-19 as a pandemic led to extensive research on understanding the virus and avenues to restrict viral infection and the spread of disease. Several exemplary research articles have ever since documented the mode of viral transmission, the rise of different mutant variants, and the various safety measures needed for protection from infection. With the marvelous effort put forward by a few companies several vaccines have also been developed. The currently ongoing vaccination drives worldwide are now aimed at controlling COVID-19. Yet, after two major waves of infection and deaths, COVID-19 persists. Moreover, much remains unknown about the level of protective immunity generated through natural infections by SARS-CoV-2, and the efficacy of vaccinations in different communities all over the globe.

We previously published the rate of SARS-Cov-2 infection and the efficacy of the Covishield vaccine in generating anti-S-RBD antibody in several pockets all over West Bengal, India [14]. In the current article we have evaluated the potential of the antibodies generated by both Covishield and Covaxin to neutralize viral infection using an in vitro assay [31], focusing on individuals in and around Kolkata, India. We have also analyzed the efficacy of Covishield and Covaxin in generating T cell mediated immunity, a major arm of immune memory that is crucial for combating viral infections [16; 17; 18; 19; 20]. Moreover, we have compared the immunity generated by vaccinations with that developed through natural infections.

As markers of cell-mediated immunity generated through vaccination or natural infection we used T cell associated IFN*γ* and CD40L, in line with the involvement of these molecules in sustaining the recall immune response generated upon antigenic stimulation of already primed T cells [16; 18; 21; 24; 26; 27]. It is to be noted, however, that IFN*γ* driven memory-T cell cytotoxicity, a major protection against viral infections can also operate independent of assistance provided by CD40L-CD40 receptor interaction [32].

In this study we demonstrated that a large fraction of the blood plasma samples collected from vaccinated individuals is effective in neutralizing the WT virus, Covishield-specific plasma being considerably more effective than Covaxin-specific plasma in this respect (Figures 1 & 2). Both groups are also effective against the DELTA virus albeit with lesser efficiency; Covisheild faring slightly better than Covaxin. Neutralizing antibody titer after natural infection (second wave), however, is markedly better than that produced by both the vaccines. The paucity of neutralizing antibodies among individuals belonging to the first wave infection group may be due to the dearth of long - lived antibody secreting plasma cells [33]. In terms of T cell mediated immunity (Figure 3, Tables 2, 3, and Supplementary Figures 4 and 5) the picture is quite different. T cell-mediated immune responses generated by the two vaccines are quite similar and in fact, Covaxin could even be considered better than Covishield in raising immune responses against the DELTA strain. Natural infection, on the other hand, not only appears to be quite similar to both the vaccines with regard to cell mediated immunity against the wild type virus, but also generates better immunity against the delta variant (Table 2, 3). Moreover, T cell mediated immunity from the first infection wave lasts for as long as ten months, indicating the persistence of memory T cells in one-time infected individuals [16; 17]. The observed lack in correlation between antibody driven immune response and cell-mediated immune response in the vaccinated groups could be on account of difference in the frequency of available antigenic epitopes for B cells and T cells in the two groups.

A few recent studies showed vaccines were significantly less protective against DELTA variant than the WT virus [34; 35]. These studies used non-pathogenic helper virus systems for the neutralization test whereas we used a purely protein-protein interaction-based assay. Both assay methods are artificial, not mimicking the true infection in a real life scenario and therefore cannot be compared. The best answer to the effectiveness of a vaccine against DELTA and other variants perhaps will come from a real time dependent population-based study. Several studies have already shown that WHO approved vaccines are quite effective against all variants currently present in the air since very few individuals have been severely sick or hospitalized after two doses of the vaccines [36; 37; 38; 39]. The trend is also similar in India [12]. Hospitalizations and deaths after vaccination have so far been associated with severe co-morbidities.

Overall, our study reveals that among the human subjects under consideration, both Covishield and Covaxin generate humoral and cell mediated immune response against WT and DELTA SARS-CoV-2. Although the vaccines vary in the nature of the immune responses generated, both are effective. Since most countries are now requiring only fully vaccinated people to travel and only WHO approved vaccines are recognized, our study provides assurance to the international vaccine authority of the positive effect of the vaccines administered in West Bengal, India. However, our study also reveals that the immune response of vaccinated people is not quite to the same level as that of the naturally infected. Whether, Covishield/Covaxin vaccinations or natural infection will generate enough long-term immunity to cross the hurdle of future infections by the SARS-CoV-2 wild type strain or its mutant variants remains to be seen.

## Data Availability

All data produced in the present work are contained in the manuscript.

## Acknowledgements

Authors thank the following nongovernment organizations and individuals whose support in sample collection was vital to complete the study: Umapati Dutta and Milan Samity, Kolkata; Subhankar Debnath, Partha Chakraborty and Prafullanagar Warriors, Kolkata; Joy Malya Deshmukh and Bokaburo, Kolkata, and the villagers of Bhadur and Gopinathpur. Authors also thank Dr. Vivien Wang, Dr. Jayanta Chowdhury, Dr. Ratna Chattopadhyay, Dr. Sandip Ghosh, Dr. Subhas Ghosh, Dr. Debi Dutta, and the entire staff of the New Avenue Nursing Home, Kolkata, for their gracious support in study organization. Tanmoy Dalui is acknowledged for his help in FACS analysis of all samples. Authors also thank the following people for assisting in subject identification and collection: Tarasankar Ghosh, Sujata Ghosh, Dipsankar Ghosh, Anushka Ghosh, Gostogopal Shee, Amal Das and Pratap Santra.

## Figure legends

Figure 1: <u>Seroreactivity of infected and vaccinated population against Spike (S)-RBD.</u> A. Seroreactivity against RBD-WT of all samples in each of the five groups. 95% confidence intervals and p values for comparing individual distributions to that healthy control are shown. B. Seroreactivity against RBD-DELTA of all samples in each of the five groups. 95% confidence intervals and p values for comparing individual distributions to that healthy control are shown.

Figure 2: <u>Competition between plasma/sera from infected and vaccinated people and ACE2 for RBD binding as surrogate neutralization activity.</u> A: Neutralization as percent signal inhibition by sera/plasma samples (1:10 dilution) from five different groups with 100 ng His-RBD-WT as the capture antigen in competition with 20 ng ACE2 by ELISA. Each sample was run in duplicates and each dot represents a mean of two readings. B: Neutralization as percent signal inhibition by the same sera/plasma samples as in A with 100 ng His-RBD-DELTA variant as the capture antigen in competition with 20 ng ACE2. Each sample was run in duplicates and each dot represents a mean of two readings. Competition assays of each sample for both RBD-WT and RBD-DELTA were done in parallel in same plate for better reproducibility. C. Competition was tested using the Ni2-NTA affinity pulldown assay. His-RBD-WT (400 ng) captured by Ni2+ affinity beads (red arrow) bound pure ACE2 (green arrow, 400 ng) (lane 1). A sample with RBD-specific antibody efficiently competed ACE2 binding (lane2) but not a sample negative for RBD-specific antibody (lane 3). ‘M’ denotes MW standards. ‘violet arrow’ denotes Ig-heavy chain that specifically bound to RBD-WT. D. Same as ‘C’ except RBD-DELTA was used as captured antigen. Positive and negative plasma used were same as in C.

Figure 3: <u>Depiction of for FACS gate for PBMC collected from human blood samples for analysis of cell mediated immunity</u>. **A**: Representation of initial gating on PBMC for CD4+ and CD8+ lymphocytes followed by demarcation of IFN*γ* and CD40L positive populations, based on evaluation of geometric mean fluorescence intensity generated by antibody staining. **B:** Representation of change in percent and geometric mean fluorescence intensity (noted in parenthesis) of IFN*γ* positive CD8T lymphocytes in response to stimulation by SARS-Cov-2 specific S-RBD protein. **C**: Similar representation with respect to CD40L positive T lymphocytes.

**Supplementary Figure 1.**
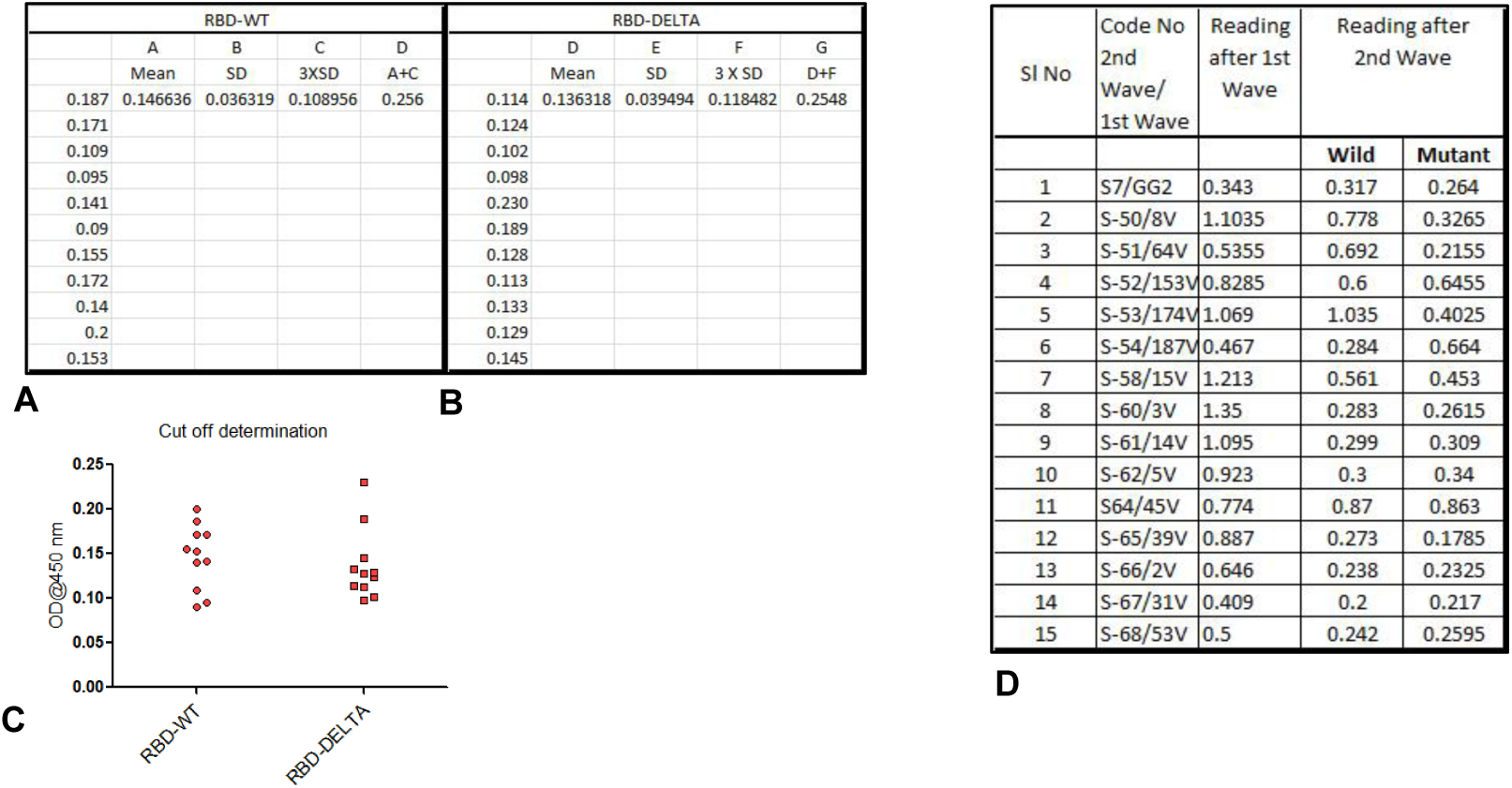
Cut off value determination using 11 negative sera samples. **A**. A_450_ reading of sera against the RBD of the spike protein of WT virus (RBD-WT) and the cut off value calculated from the mean and standard deviation. **B**. A_450_ reading of sera against the S-RBD-DELTA and the cut off value. **C**. A_450_ distribution plot using the program graphpad. **D**. Comparison of antibody titer against S-RBD for the 15 first wave samples taken six months apart (January 2021) and (June 2021)

**Supplementary Figure 2.**
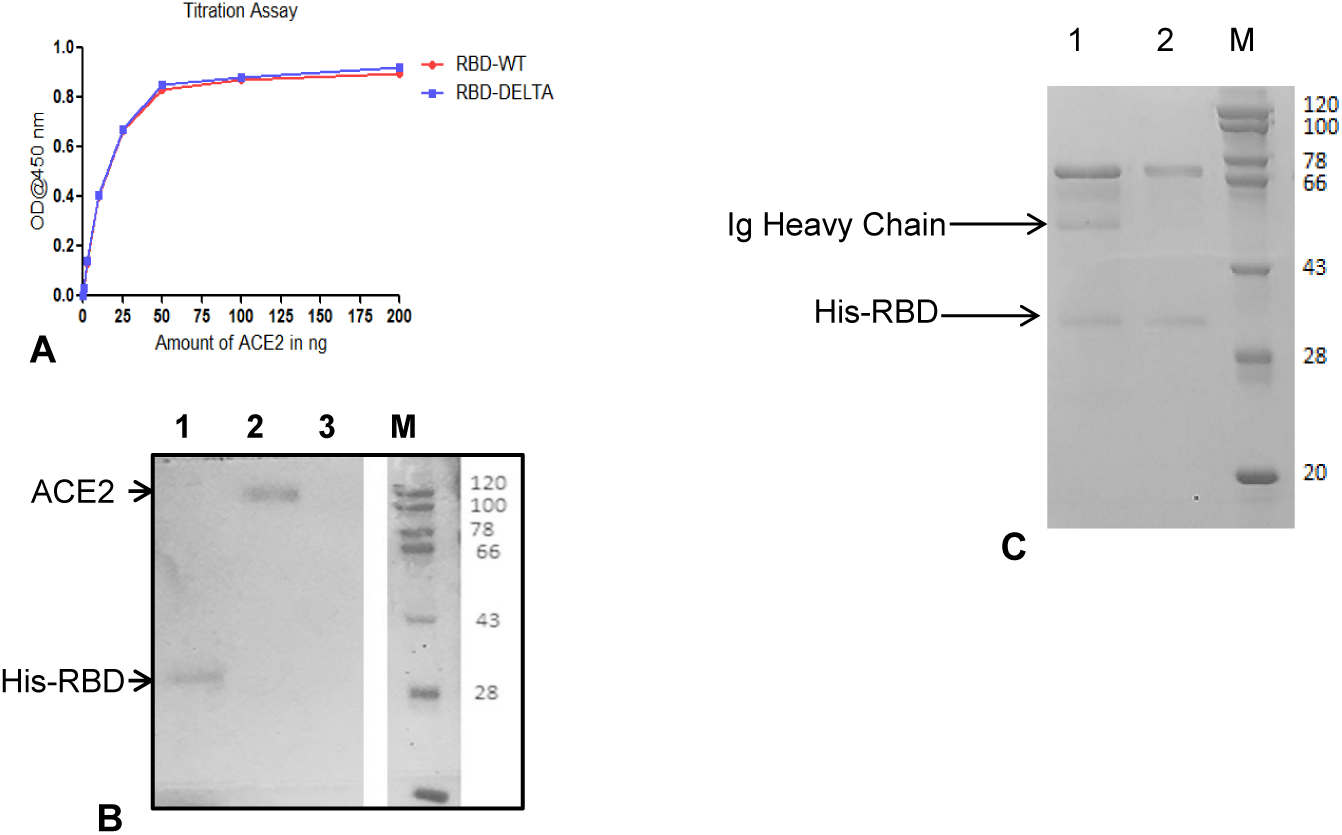
Interactions between S-RBD and ACE2 or Specific Ig. **A**. ELISA-based binding titration where increasing concentrations of ACE2 protein was added to wells coated with 100 ng RBD-WT (red) or RBD-DELTA. RBD proteins were prepared as poly-histidine fusion proteins and ACE2 was prepared as a Fc fusion protein. **B**. Binding interaction between His-RBD-WT and ACE2 showing His-pulldown assay followed by SDS-PAGE separation and staining with Coommassie blue. Lane 1: His-RBD incubated with Ni-NTA beads. Lane 2: ACE2 incubated with beads bound to His-RBD-WT, Lane 3: ACE2 incubated with Ni-NTA beads (control). M denotes MW standard. **C**. Binding interaction between His-RBD-WT and Spike (S) protein-specific antibody in plasma or control plasma by His-pulldown assay followed by SDS-PAGE separation and staining with Coommassie blue. His-RBD-WT pulled down heavy chain of specific antibody (IgG) (lane 1) but no IgG heavy could be seen in seronegative sample (lane 2).

**Supplementary Fig 3.**
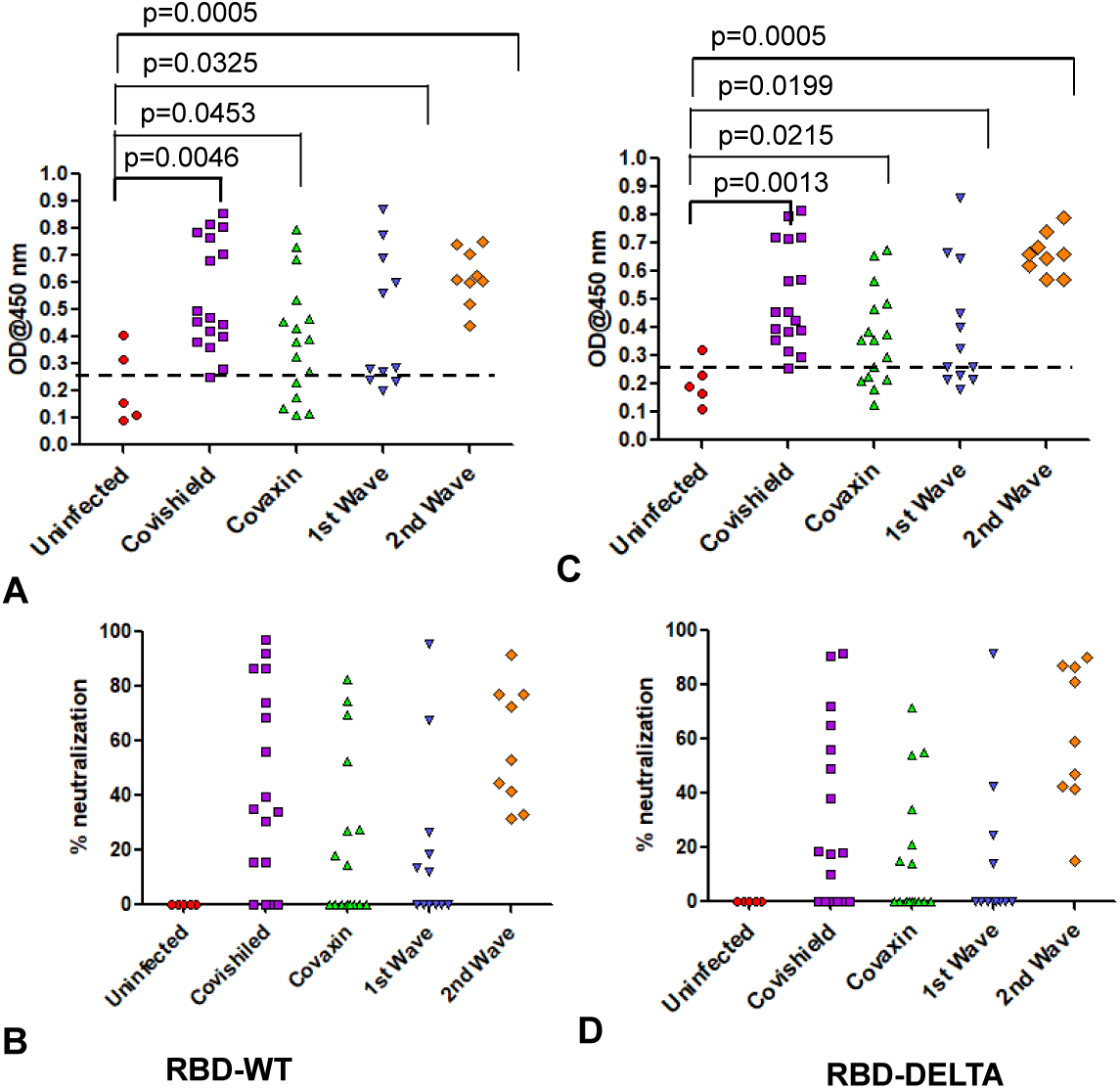
Humoral immunity of samples of all five groups used for cell mediated immunity. **A** & **B:** Humoral immunity against RBD-WT. Seropositivity (A) and neutralization efficiency (B) against RBD-WT. **C** & **D:** Humoral immunity against RBD-DELTA. Seropositivity (C) and neutralization efficiency (D) against RBD-DELTA.

**Supplementary Figure 4.**
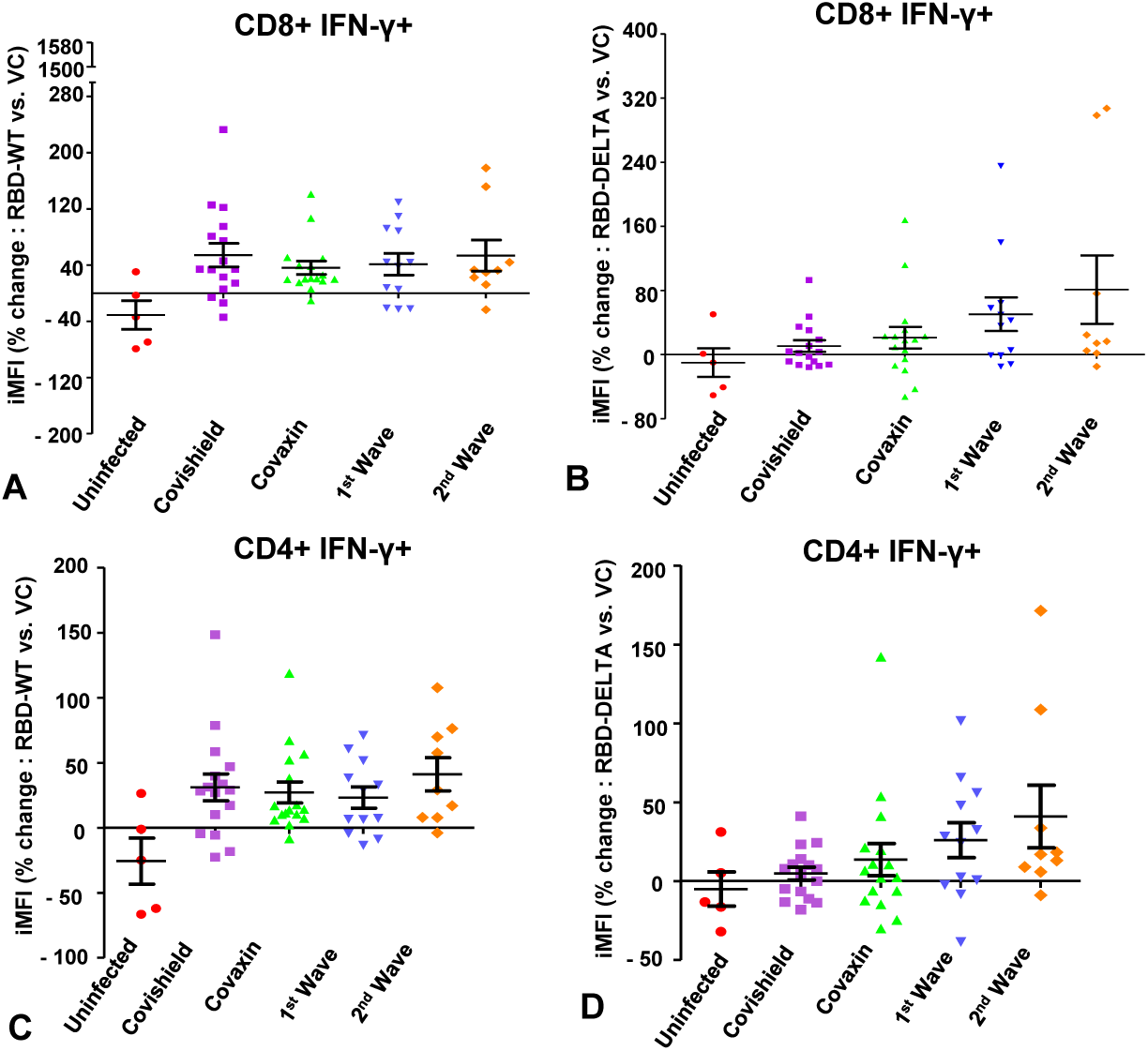
Evaluation of immune response to wild type and mutant S-RBD protein with respect to IFN*γ* in CD8+ and CD4+ T lymphocytes. **A & B**: SARS Cov-2-S-RBD (A: Wild type S-RBD, B: Mutant S-RBD) specific percent change in iMFI (RBD vs. vehicle control:vc) of IFN*γ* in CD8 T lymphocytes of individuals belonging to 5 different groups: (i) uninfected, (ii) vaccinated with Covishield, (iii) vaccinated with Covaxin, (iv) first wave infection and (v) second wave infection. **C & D**: SARS Cov-2 – S-RBD (C: Wild type, D: Delta) specific percent change in iMFI of IFN*γ* in CD4 T lymphocytes of individuals belonging to the same groups as summarized in A & B.

**Supplementary Figure 5.**
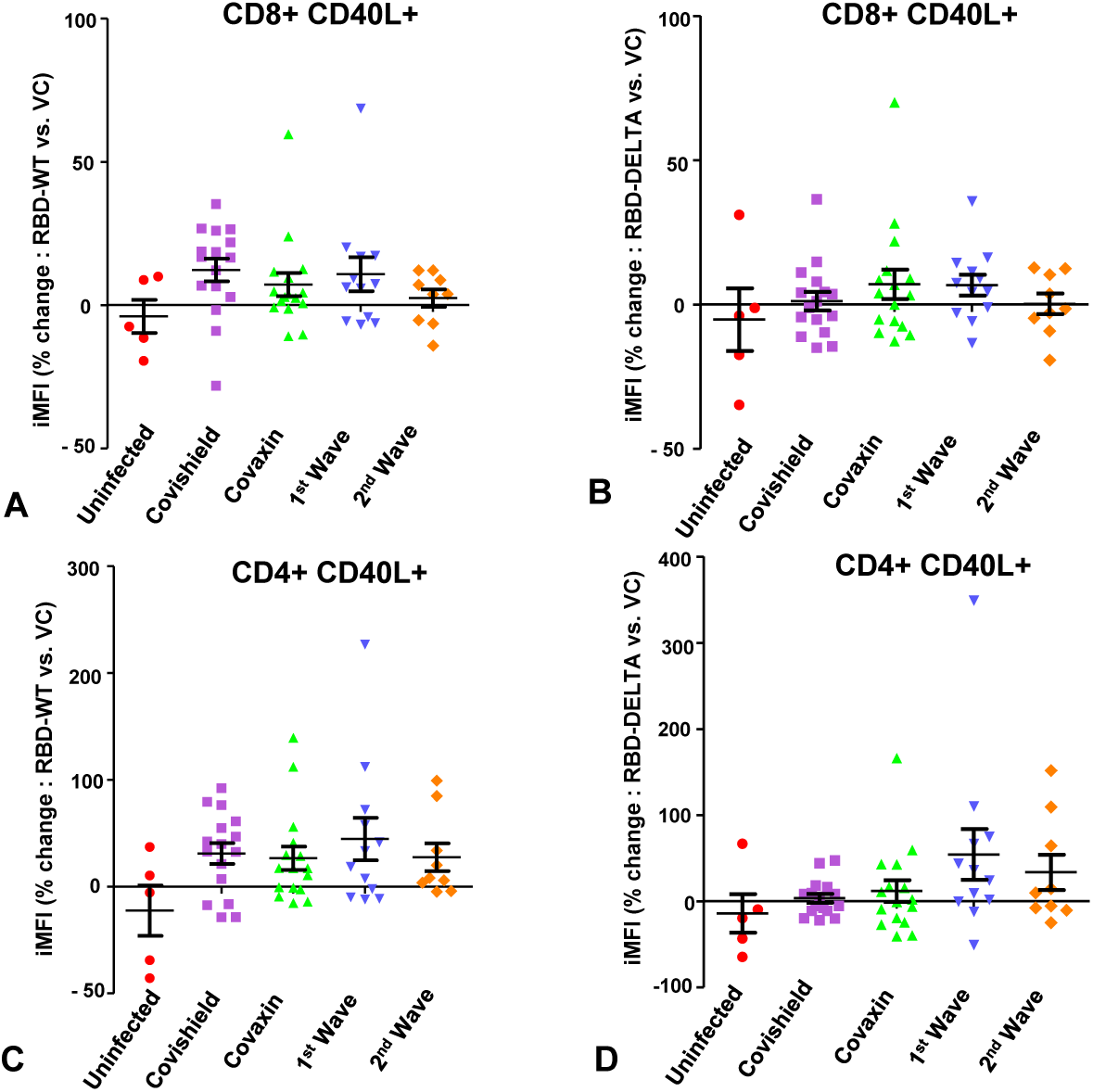
**A & B**: S-RBD (A: Wild type, B: DELTA) specific percent change in iMFI of CD40L (RBD vs. vehicle control:vc in CD8 T lymphocytes. **C & D**: Similar representation of S-RBD specific (C: Wild type, D: Delta) change in CD40L expression in CD4 T cells.

**Supplementary Table 1.** Summary of test subjects for assessment of cell mediated immunity

| **Groups** | **Number of Subjects** |
| --- | --- |
| Covishield | 16 |
| Covaxin | 16 |
| Infection 1st wave | 12 |
| Infection 2nd wave | 9 |
| Uninfected | 5 |

